# Fesoterodine ameliorates autonomic dysreflexia while improving lower urinary tract function and urinary incontinence-related quality of life in individuals with spinal cord injury: A prospective phase IIa study

**DOI:** 10.1101/2022.07.14.22277625

**Authors:** Matthias Walter, Andrea L. Ramirez, Amanda H. X. Lee, Thomas E. Nightingale, Daniel Rapoport, Alex Kavanagh, Andrei V. Krassioukov

## Abstract

The aim of this prospective phase IIa, open-label exploratory, pre-post study was to determine the efficacy of fesoterodine to ameliorate autonomic dysreflexia (AD) in individuals with chronic SCI (>1-year post-injury) at or above the sixth thoracic spinal segment, with confirmed history of AD and neurogenic detrusor overactivity (NDO). We screened 20 individuals. Fifteen individuals provided written informed consent and were assigned to undergo urodynamics, 24-hour ambulatory-blood-pressure-monitoring (ABPM), and urinary incontinence-related quality of life (QoL) measures at baseline and on-treatment. The Montreal Cognitive Assessment (MoCA) and Neurogenic Bowel Dysfunction (NBD) score were used to monitor cognitive and bowel function, respectively. Twelve participants (4 females, median age 42 years) completed this study. Compared to baseline, fesoterodine improved lower urinary tract (LUT) function, i.e., increased cystometric capacity (205 vs 475mL, p = 0.002) and decreased maximum detrusor pressure (44 vs 12cmH_2_O, p = 0.009). NDO was eliminated in seven (58%) participants. Severity of AD events during urodynamics (40 vs 27mmHg, p = 0.08) and 24-hour ABPM (59 vs. 36mmHg, p = 0.05) were both reduced, yielding a large effect size (*r* = -0.58). AD Frequency (14 vs. 3, p = 0.004) during 24-hour ABPM was significantly reduced. Urinary incontinence-related QoL improved (68 vs. 82, p = 0.02), however, cognitive (p = 0.2) and bowel function (p = 0.4) did not change significantly. In conclusion, fesoterodine reduces the magnitude and frequency of AD, while improving LUT function and urinary incontinence-related QoL in individuals with chronic SCI without negatively affecting cognitive or bowel function.

## INTRODUCTION

Neurogenic detrusor overactivity (NDO) and autonomic dysreflexia (AD) combine to place a tremendous burden on health and quality of life (QoL) in individuals living with a spinal cord injury (SCI). We have previously shown that the presence of NDO and the neurological level of injury (NLI) are independent risk factors for developing AD during urodynamic studies (UDS).^1^ The higher the NLI above the sixth thoracic spinal cord segment (T6), the higher the odds of experiencing AD. Since AD can lead to potentially life-threatening complications, such as stroke, myocardial infarction, or even death, urologists should take precautions when conducting UDS in this population.^2^ Furthermore, we have provided evidence that onabotulinumtoxinA, a second-line treatment option, ameliorates AD while effectively improving lower urinary tract (LUT) function and urinary incontinence-related QoL.^3^ However, whether antimuscarinics (i.e., first-line treatment option) have the capacity to ameliorate AD in this cohort has not yet been investigated. Thus, our aim was to determine whether fesoterodine is effective in reducing the incidence and severity of AD episodes during UDS and in daily life in individuals with chronic (>1-year post-injury) SCI ≥ T6.^4^

## MATERIAL AND METHODS

This prospective phase IIa, open-label exploratory, non-blinded, non-randomised, single-centre pre-post study was approved by the University of British Columbia Clinical Research Ethics Board (H15-02364), Vancouver Coastal Health Research Institute (V15-02364) and Health Canada (205857). Furthermore, this study was registered at clinicaltrials.gov (identifier NCT02676154). A study protocol, adhering to the standard protocol items: recommendations for interventional trials and consolidated standards of reporting trials statements has been previously published.^4^

After screening twenty individuals with chronic SCI ≥ T6, fifteen individuals with confirmed history of AD and NDO provided written informed consent according to the Helsinki II declaration and underwent a battery of baseline assessments (Figure 1). The NLI and completeness (i.e. American Spinal Injury Association impairment scale [AIS] grade) of SCI were classified according to the International Standards for Neurological Classification of SCI.^5^ All UDS (Aquarius TT, Laborie Model 94-R03-BT, Montreal, Quebec, Canada) were performed in accordance with the International Continence Society.^6^ Concurrent to UDS, we continuously recorded beat-by-beat blood pressure via finger photoplethysmography (Finometer PRO, Finapres Medical Systems, Amsterdam, Netherlands), corrected to brachial pressure (CARESCAPE V100, GE Healthcare, Milwaukee, WI, USA), and one-lead electrocardiogram (eML 132; ADInstruments, Colorado Springs, CO, USA) for heart rate in order to detect AD.^1, 7^

**Figure 1.**
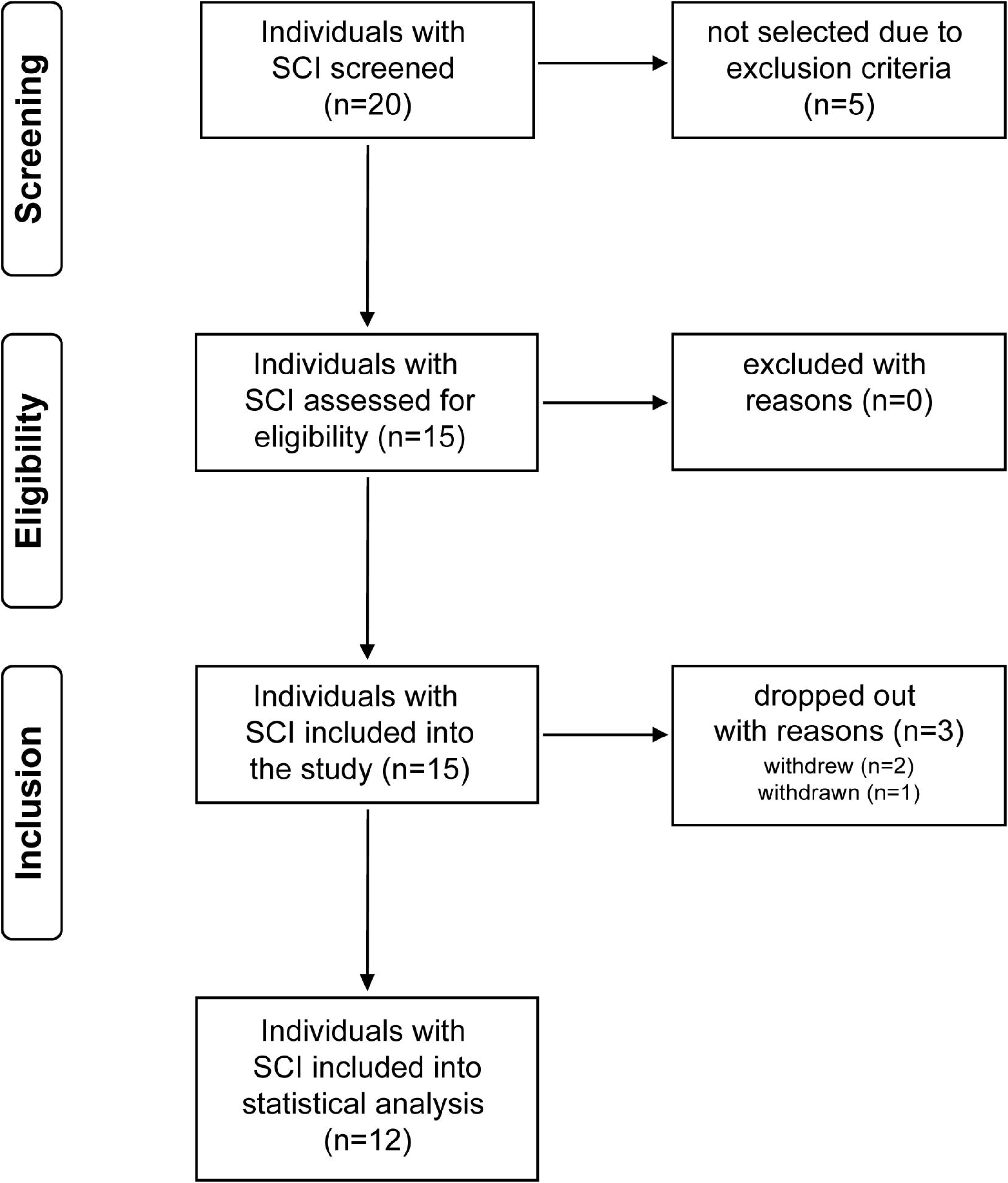
Study flow diagram.

After confirmation of AD during baseline UDS, frequency and severity of AD in daily life were recorded using 24-hour ambulatory-blood-pressure-monitoring (ABPM, Meditech Card(X)plore device, Meditech, Budapest, Hungary).^8^ All participants completed validated, standardized questionnaires to subjectively monitor urinary incontinence-related QoL (I-QoL),^9^ AD health-related QoL (AD-HR-QoL)^8^, bowel function (neurogenic bowel dysfunction [NBD] Score)^10^ and cognitive function (Montreal cognitive assessment [MoCA]^11^), respectively. Ten to twelve weeks following the start of treatment, objective and subjective measures were repeated to assess on-treatment efficacy.

The aim of this study was to assess the effect of fesoterodine (i.e., 12-week treatment period; on-treatment compared to baseline) in reducing the severity of AD (i.e., maximum increase in systolic blood pressure [SBP]) during UDS, as well as severity and frequency of AD occurring in daily living as detected during the 24-hour ABPM. The two primary outcome measures were number of participants who experienced a decrease in severity of AD during UDS and 24-hour ABPM. Secondary outcome measures included: the improvement in UDS parameters (e.g., cystometric capacity and detrusor pressure); number of participants who experienced a decrease in the frequency of AD in daily life (i.e., during 24-hour ABPM); number of participants who experienced a reduction in self-reported AD severity and frequency (i.e., AD-HR-QoL); an improvement of self-reported urinary incontinence-related QoL (i.e., I-QoL); an improvement in bowel (i.e., NBD Score) and cognitive function (i.e., MoCA, total score ≥26 considered as unimpaired cognitive function).

Following baseline assessments, eligible individuals received a 4-week supply of 4mg daily doses of fesoterodine. During the treatment period, individuals returned to the study centre (i.e., at the latest 2 days before their supply ran out). During these visits, participants were assessed for dose efficacy. In consultation with the investigator, individuals had a choice to either increase the dose of the study drug to 8mg or maintain the same dose (4mg). Participants who elected to increase their dose to 8mg per day had the option to return to 4mg at any time. However, participants only had the option to increase their dose once, meaning that no further increase in dose was permitted following a dose reduction. Study drug compliance was monitored using a diary to identify missed doses. Participants were asked to indicate the days where doses were missed. Non-adherence was considered when an individual failed to take fesoterodine consecutively (>5 days) or intermittent (>50% of all days within one cycle). Lastly, we recorded any adverse drug reactions (ADRs) over the course of the 12-week treatment period.

Statistical analysis was performed using R Statistical Software Version 4.0.5 for Mac Os. Considering the limited size of our cohort, non-parametric statistics (i.e. Wilcoxon signed-rank test) were used to compare within participants (i.e. baseline vs. on-treatment assessment). Data are presented as median with lower and upper quartiles (Q1; Q3); and minimum and maximum for age and time post-injury). Furthermore, effect size expressed as Pearson correlation coefficient, i.e. Pearson’s (*r*) was calculated as Z statistics divided by square root of total number of pairs (N) in accordance with Rosenthal:^12^

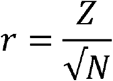

Pearson’s *r* can vary in magnitude from -1 to 1, with -1 indicating a perfect negative linear relation, 1 indicating a perfect positive linear relation, and 0 indicating no linear relation between two variables (effect sizes: small, *r* = 0.1 – 0.29 or -0.1 – (−0.29); medium, *r* = 0.3 – 0.49 or -0.3 – (−0.49); large, *r* ≥ 0.5 or -0.5).

## RESULTS

In total, 12 individuals [4 females (33 %), mean age 42 years (36; 50, 29 – 52) and mean time post-injury 19 years (12; 22, 7 – 39)] completed the study and were included for analysis (Table 1). The majority had cervical (n=8), motor-complete (AIS A/B = 10) SCI.

**Table 1.**
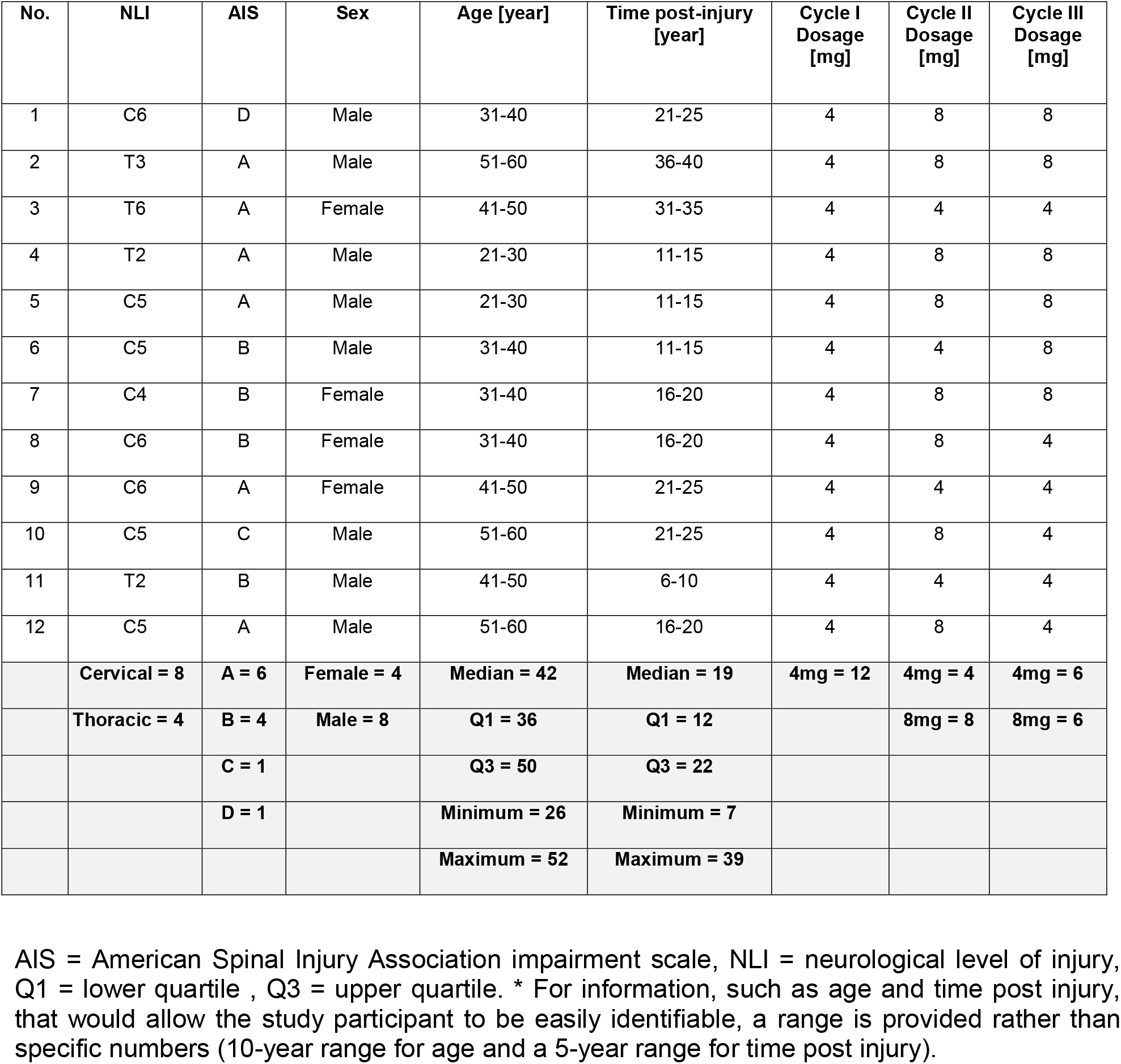
Participant characteristics.

| No. | NLI | AIS | Sex | Age [year] | Time post-injury [year] | Cycle I Dosage [mg] | Cycle II Dosage [mg] | Cycle III Dosage [mg] |
| --- | --- | --- | --- | --- | --- | --- | --- | --- |
| 1 | C6 | D | Male | 31-40 | 21-25 | 4 | 8 | 8 |
| 2 | T3 | A | Male | 51-60 | 36-40 | 4 | 8 | 8 |
| 3 | T6 | A | Female | 41-50 | 31-35 | 4 | 4 | 4 |
| 4 | T2 | A | Male | 21-30 | 11-15 | 4 | 8 | 8 |
| 5 | C5 | A | Male | 21-30 | 11-15 | 4 | 8 | 8 |
| 6 | C5 | B | Male | 31-40 | 11-15 | 4 | 4 | 8 |
| 7 | C4 | B | Female | 31-40 | 16-20 | 4 | 8 | 8 |
| 8 | C6 | B | Female | 31-40 | 16-20 | 4 | 8 | 4 |
| 9 | C6 | A | Female | 41-50 | 21-25 | 4 | 4 | 4 |
| 10 | C5 | C | Male | 51-60 | 21-25 | 4 | 8 | 4 |
| 11 | T2 | B | Male | 41-50 | 6-10 | 4 | 4 | 4 |
| 12 | C5 | A | Male | 51-60 | 16-20 | 4 | 8 | 4 |
|  | **Cervical = 8** | **A = 6** | **Female = 4** | **Median = 42** | **Median = 19** | **4mg = 12** | **4mg = 4** | **4mg = 6** |
|  | **Thoracic = 4** | **B = 4** | **Male = 8** | **Q1 = 36** | **Q1 = 12** |  | **8mg = 8** | **8mg = 6** |
|  |  | **C = 1** |  | **Q3 = 50** | **Q3 = 22** |  |  |  |
|  |  | **D = 1** |  | **Minimum = 26** | **Minimum = 7** |  |  |  |
|  |  |  |  | **Maximum = 52** | **Maximum = 39** |  |  |  |

AIS = American Spinal Injury Association impairment scale, NLI = neurological level of injury, Q1 = lower quartile , Q3 = upper quartile. \* For information, such as age and time post injury, that would allow the study participant to be easily identifiable, a range is provided rather than specific numbers (10-year range for age and a 5-year range for time post injury).

Regarding our primary outcome, 10 (83%) and 9 (75%) participants experienced a decrease in severity of AD during UDS and during daily life, respectively. Further, fesoterodine ameliorated objectively measured AD, i.e., smaller increase (Δ) in systolic blood pressure (SBP) during on-treatment UDS compared to baseline [Figure 2A, 40 mmHg (24; 44) vs. 27 mmHg (14; 33), p = 0.08, Z = -2, *r* = -0.58] and severity of AD (ΔSBP) until cystometric capacity from the baseline UDS was reached during on-treatment UDS [Figure 2B, 40 mmHg (24; 44) vs. 4.5 mmHg (0; 10.5), p = 0.002, Z = -3, *r* = -0.87]. Furthermore, the severity [Figure 2C, 59 mmHg (48; 69) vs. 36 mmHg (28; 56), p = 0.04, Z = -2, *r* = -0.58] and frequency [Figure 2D, 14 (5; 28) vs. 3 (2; 12), p = 0.004, Z = -3, *r* = -0.87], of AD during daily life measured by 24-h-ABPM were significantly reduced on-treatment. Subjectively, fesoterodine reduced the frequency [Figure 2E, 8.5 (6; 11) vs. 7 (4.2; 9.2), p = 0.2, Z = -1, *r* = -0.29] and severity [Figure 2F, 4.5 (2.8; 8.5) vs. 3 (2; 6.5), p = 0.2, Z = -1, *r* = -0.29] of bladder-related AD symptoms in daily life.

**Figure 2.**
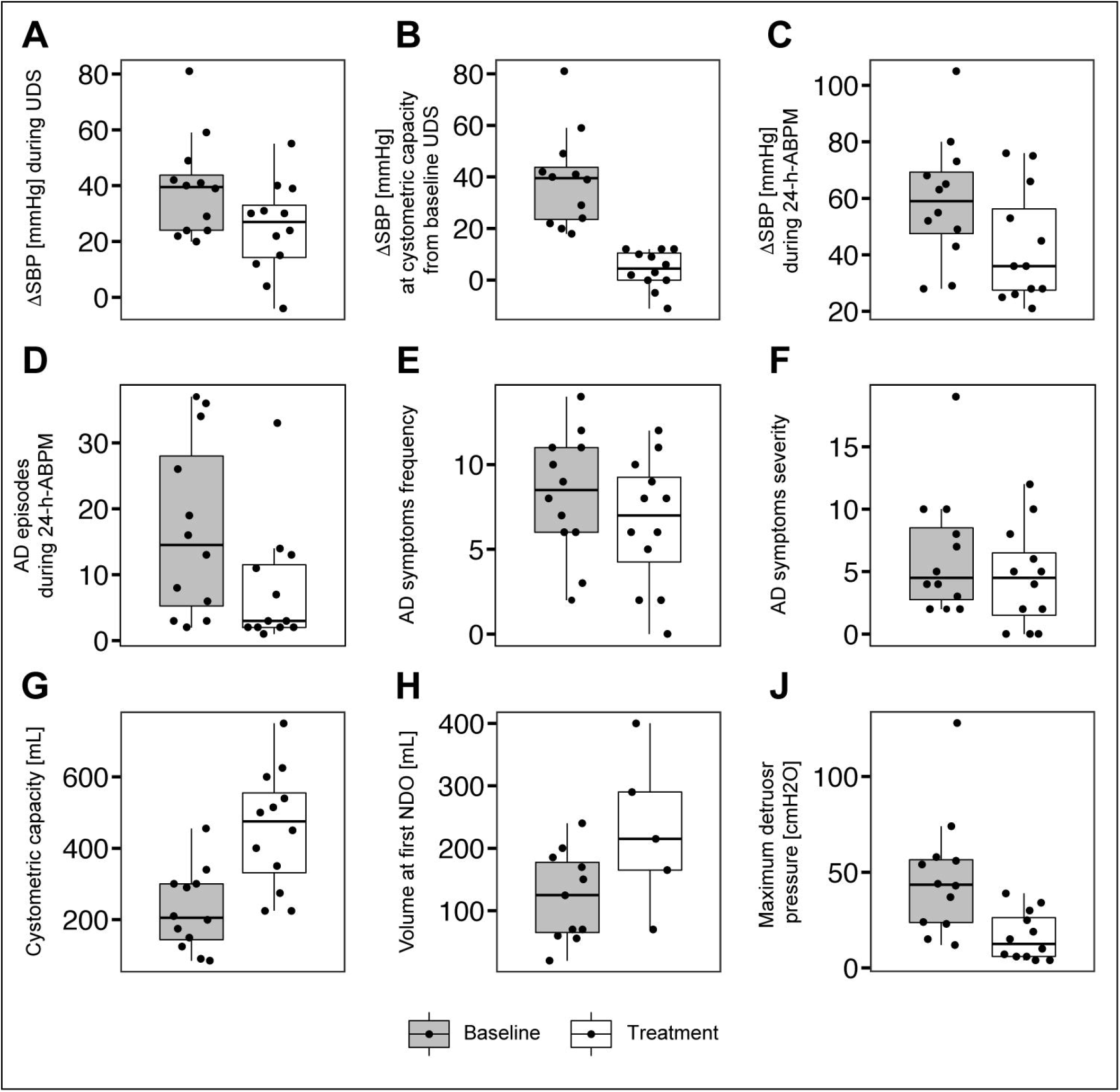
Effect of fesoterodine on AD during UDS and in daily life, and on LUT function: Comparison between on-treatment and baseline assessments for (A) severity of AD (i.e. increase in systolic blood pressure, ΔSBP) during UDS, (B)* severity of AD until cystometric capacity from baseline UDS was reached during on-treatment UDS, (C)* severity of AD in daily life (i.e. during 24-h ABPM), (D)* frequency of AD episodes in daily life, (E) AD symptoms frequency score, (F) AD symptoms severity score, (G)* cystometric capacity, (H) volume at first NDO, and (J)* maximum detrusor pressure during bladder filling (i.e. storage). *Data are presented at group level using boxplots (median, interquartile range) and individually (dots)*. *\* Statistically significant changes (p<0*.*05)*

Further, fesoterodine objectively improved LUT function. Cystometric capacity [Figure 2G, 205 mL (144; 300) vs. 475 mL (331; 555), p = 0.002, Z = 3, *r* = 0.87] increased significantly. Volume at first NDO [Figure 2H, 125 mL (65; 178) vs. 215 mL (165; 290), p = 0.1, Z = 2, *r* = 0.58] also increased but did not yield statistical significance. However, the effect of volume increase was large, considering that only five individuals (−58%) had NDO while being on-treatment. Further, fesoterodine significantly decreased maximum detrusor pressure during bladder filling [Figure 2J, 44 cmH_2_O (24; 56) vs. 12 cmH_2_O (6; 26), p = 0.009, Z = -3, *r* = -0.87].

In addition, urinary incontinence-related QoL, assessed using the I-QoL questionnaire, was significantly improved overall, i.e., *in total* [Figure 3A, 68 (55; 80) vs. 82 (77; 90), p = 0.02, Z = 2, *r* = 0.58] as well as in sub-categories *Psychological Impact* [Figure 3B, 84 (54; 95) vs. 92 (83; 100), p = 0.006, Z = 3, *r* = 0.87] and *Social Embarrassment* [Figure 3C, 50 (39; 80) vs. 78 (55; 90), p = 0.04, Z = 2, *r* = 0.58]. In addition, sub-category *Avoidance* [Figure 3D, 68 (50; 84) vs. 82 (77; 88), p = 0.1, Z = 2, *r* = 0.58] was improved by a large magnitude but did not yield statistical significance. Further, we observed no changes in bowel function, i.e. NBD total score [Figure 3E, 9.0 (6.0; 12.5) vs. 8.5 (6.0; 13.2), p = 0.7, Z = 0, *r* = 0; and NBD general satisfaction [7 (5.8; 8) vs. 8 (5.8; 8), p = 0.4, Z = 1, *r* = 0.29], without any negative effect on cognitive function [Figure 3F, MoCA, 29.0 (25.8; 29.2) vs. 29.0 (28.0; 30), p = 0.2, Z = 1, *r* = 0.29].

**Figure 3.**
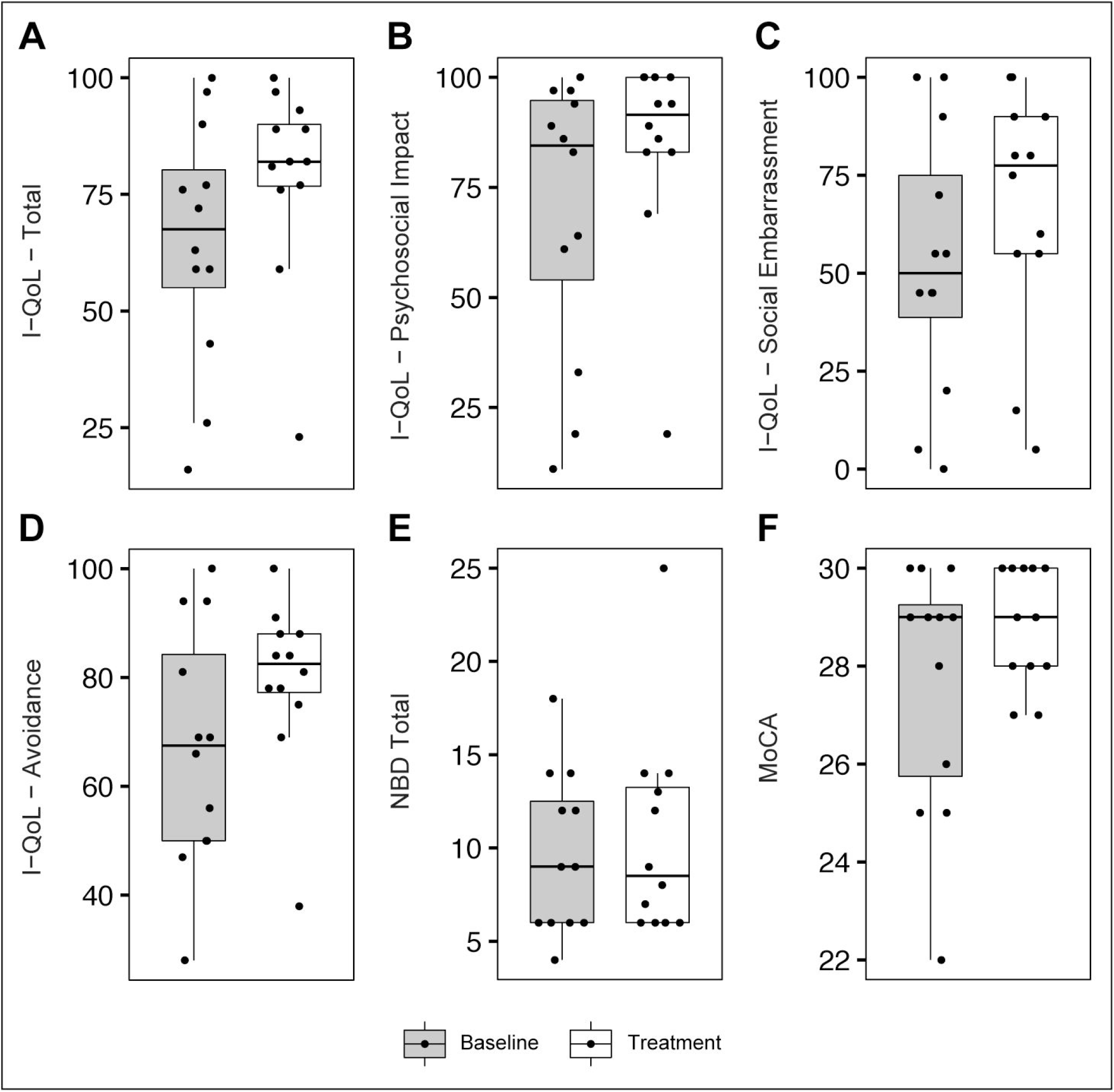
Effect of fesoterodine on urinary incontinence-related QoL, bowel and cognitive function: Comparison between on-treatment and baseline assessments for urinary incontinence related QoL, i.e., I-QoL (A)* *Total*, with subcategories (B)* *psychosocial impact*, (C)* *social embarrassment*, and (D) *avoidance and limiting behavior* as well as bowel, i.e. (E) *NBD* score and cognitive function, i.e. (F) *MoCA*. *Data are presented at group level using boxplots (median, interquartile range) and individually (dots). * Statistically significant changes (p<0*.*05)*

All 12 participants adhered to the study protocol including the intake of fesoterodine. At the end of the treatment phase, daily dosage distribution among participants was even, i.e. 4mg (n = 6) or 8mg (n = 6). Overall, we recorded 26 ADRs in 10 participants (Table 2), i.e. related (n = 23) or possibly related (n = 3), which were all grade 1 (n = 21) or 2 (n = 5).

**Table 2.** Safety monitoring highlighting the number and distribution of adverse drug reactions.

| Adverse drug reactions (ADRs) | Overall frequency | ADRs per 4-week cycle and Fesoterodine dosage |  |  |
| --- | --- | --- | --- | --- |
|  |  | I (4mg) | II (4 or 8mg) | III (4 or 8mg) |
| **Related\*** | **23 (88%)** | **15** | **14 (3 / 11)** | **15 (10 / 5)** |
| Dry mouth | 9 | 7 | 5 (1 / 4) | 5 (3 / 2) |
| Dry eyes | 3 | 2 | 1 (0 / 1) | 2 (2 / 0) |
| Fatigue | 3 | 2 | 3 (1 / 2) | 3 (1 / 2) |
| Increased constipation | 2 | 0 | 2 (0 / 2) | 2 (1 / 1) |
| Dyspepsia | 2 | 0 | 2 (0 / 2) | 2 (2 / 0) |
| Increased GGT level | 1 | 1 | 1 (1 / 0) | 1 (1 / 0) |
| Dry skin | 1 | 1 | 0 | 0 |
| Dizziness | 1 | 1 | 0 | 0 |
| Somnolence | 1 | 1 | 0 | 0 |
| **Possibly related** | **3 (12%)** | **3** | **2 (0 / 2)** | **2 (1 / 1)** |
| Decreased libido | 1 | 1 | 1 (0 / 1) | 1 (0 / 1) |
| Reduced sensation of touch | 1 | 1 | 0 | 0 |
| Fecal incontinence | 1 | 1 | 1 (0 / 1) | 1 (1 / 0) |

\* Indicating known adverse drug reactions; GGT = Gamma-glutamyl transferase

## DISCUSSION

The majority of our cohort experienced a decrease in severity of AD during UDS and in daily life without any significant deterioration of cognitive or bowel function. Further, in line with our previous study, highlighting an efficacious second-line treatment (i.e. intradetrusor onabotulinumtoxinA injections),^3^ we observed significant improvements of LUT function and urinary incontinence-related QoL in individuals being on-treatment with fesoterodine.

Yonguc et al.^13^ reported significant improvements in overactive bladder (OAB) symptoms in older patients with Parkinson’s disease (i.e., mean age 66 years) on-treatment with fesoterodine 4mg without affecting cognitive function. In another study, DuBeau et al.^14^ showed that fesoterodine (i.e., 12-week treatment 4mg to 8mg per day) not only led to significantly greater improvements in urgency urinary incontinence episodes per 24 hours and QoL in the elderly (i.e., mean age 75 years) but also did not negatively affect cognitive function (i.e., mini-mental state examination) compared to placebo. Fesoterodine is the only antimuscarinic agent with a ‘fit for the aged’ (FORTA) classification B (i.e. beneficial, “*drugs with proven or obvious efficacy in older people, but limited extent of effect or safety concerns*”).^15^ Wagg et al.^16^ also highlighted the clinical efficacy and safety of OAB treatment (i.e., 12 weeks with 4mg to 8mg per day) in patients aged ≥65 years. Although our cohort was younger than the aforementioned studies, i.e., <65 years of age, our findings confirm the previously established safety profile of fesoterodine (i.e., only grade 1 and 2 ADRs). Further, we did not observe a dosage-dependent frequency or distribution of ADRs.

Given the vulnerability of our cohort with respect to cognitive impairment,^17^ these findings are important, as fesoterodine (as well as other antimuscarinics) is not only a first-line treatment option but for some individuals is the only option covered by their healthcare insurance. For example, Canadian provincial healthcare coverage often does not include second-line treatments, such as onabutulinumtoxinA, thus presenting significant socioeconomic burden. Given its design, our study has several limitations, such as a lack of blinding, placebo group, and follow-up beyond 3 months, which should be considered when interpreting our findings.

## CONCLUSIONS

In conclusion, our findings highlight that fesoterodine, a first-line treatment option for NDO, ameliorates AD during UDS and in daily life in individuals with SCI ≥ T6. Fesoterodine also improves LUT function and urinary incontinence-related QoL without negatively affecting bowel and cognitive function. Considering the increased risk of cardiovascular disease in this cohort,^18^ these findings are crucial as sudden increases in systolic blood pressure can result in life-threatening consequences, jeopardizing the well-being and QoL of individuals with SCI.

## Supporting information

Supplemental - Strobe checklist

## Data Availability

All data produced in the present study are available upon reasonable request to the authors.

## Author contributions

Andrei V. Krassioukov had full access to all the data in the study and takes responsibility for the integrity of the data and the accuracy of the data analysis.

## Study concept and design

Matthias Walter, Andrea Ramirez, Daniel Rapoport, Alex Kavanagh, Andrei V. Krassioukov

## Acquisition of data

Matthias Walter, Andrea Ramirez, Amanda H.X. Lee, Thomas E. Nightingale, Daniel Rapoport, Alex Kavanagh, Andrei V. Krassioukov

## Statistical analysis

Matthias Walter

## Drafting of the manuscript

Matthias Walter

## Critical revision of the manuscript for important intellectual content

Andrea Ramirez, Amanda H.X. Lee, Thomas E. Nightingale, Daniel Rapoport, Alex Kavanagh, Andrei V. Krassioukov

## Obtaining funding

This study was conducted as an investigator-initiated phase IIa clinical trial funded by Pfizer Canada (WI207218, PI. Dr Krassioukov). Matthias Walter (Postdoctoral Research Trainee Award from the Michael Smith Foundation for Health Research in partnership with the Rick Hansen Foundation under grant number 17110). Tom E. Nightingale (Postdoctoral Research Trainee Award from the Michael Smith Foundation for Health Research in partnership with the International Collaboration On Repair Discoveries under grant number grant number 17767). Dr. Krassioukov is supported by Endowed Chair in Rehabilitation, ICORD, University of British Columbia.

## Administrative, technical, or material support

Teresa Lim, Grace Coo, Ivy Allard, Colleen McLean, Tammy Wilder

## Supervision

Andrei V. Krassioukov

